# Detection of hemagglutinin H5 influenza A virus sequence in municipal wastewater solids at wastewater treatment plants with increases in influenza A in spring, 2024

**DOI:** 10.1101/2024.04.26.24306409

**Authors:** Marlene K. Wolfe, Dorothea Duong, Bridgette Shelden, Elana M. G. Chan, Vikram Chan-Herur, Stephen Hilton, Abigail Harvey Paulos, Alessandro Zulli, Bradley J. White, Alexandria B. Boehm

## Abstract

Prospective influenza A (IAV) RNA monitoring at 190 wastewater treatment plants across the US identified increases in IAV RNA concentrations at 59 plants in spring 2024, after the typical seasonal influenza period, coincident with the identification of highly pathogenic avian influenza (subtype H5N1) circulating in dairy cattle in the US. We developed and validated a hydrolysis-probe RT-PCR assay for quantification of the H5 hemagglutinin gene. We applied it retrospectively to samples from three plants where springtime increases were identified. The H5 marker was detected at all three plants coinciding with the increases. Plants were located in a state with confirmed outbreaks of highly pathogenic avian influenza, H5N1 clade 2.3.4.4b, in dairy cattle. Concentrations of the H5 gene approached overall influenza A virus gene concentrations, suggesting a large fraction of IAV inputs were H5 subtypes. At two of the wastewater plants, industrial discharges containing animal waste, including milk byproducts, were permitted to discharge into sewers. Our findings demonstrate wastewater monitoring can detect animal-associated influenza contributions, and highlight the need to consider industrial and agricultural inputs into wastewater. This work illustrates wastewater monitoring’s value for comprehensive influenza surveillance for diseases with zoonotic potential across human and animal populations.

## Introduction

Respiratory infections are the fourth largest cause of death worldwide^1–3^. Among these, influenza viruses are one of the most common causes of viral epidemics of respiratory infection, with an estimated one billion cases each year^4^. In recent years, a majority of these influenza infections have been caused by Influenza A virus (IAV)^5^, which has been responsible for all global epidemics of influenza. Influenza A viruses are found in both humans and animals; H5N1 influenza is a highly pathogenic avian influenza (HPAI) that emerged in the late 1990s and has become widespread in wild birds and has zoonotic potential. A new H5N1 clade (2.3.4.4b) has caused widespread outbreaks since 2022^6^. Tens of millions of chickens have been culled in the US to control outbreaks, and there is a threat of zoonotic transmission to humans. In 2024 this outbreak spread to dairy cattle and was associated with one known human case, the second case of HPAI documented in the United States^6^.

Wastewater surveillance of influenza A has been implemented across the United States, and concentrations of viral RNA in wastewater have been shown to align with clinical surveillance metrics including influenza-like illness, influenza hospitalizations, and deaths.^7–10^ As a result, it is becoming a routine public health tool for influenza outbreaks and provides low-cost, real-time, population level data on influenza outbreaks, particularly in locations with limited clinical testing^11^. The utility of these data and responses are predicated on the assumption that the majority of the contributions to these measurements are from human sources, rather than influenzas with zoonotic potential such as H5N1 and H7N9^12,13^. Recent increases in the spread of HPAI to new species, particularly subtype H5N1 clade 2.3. 4.4b spread in dairy cattle^6^, have raised questions about wastewater contributions from avian and bovine sources^14,15^.

Studies have demonstrated the potential for subtyping circulating IAV directly from wastewater samples^7,16^. IAV subtypes are differentiated by two main surface glycoproteins, hemagglutinin (HA) and the neuraminidase (NA)^17^. We previously applied hydrolysis probe-based RT-PCR assays for H1, H3, N1, and N2 to wastewater to identify circulating IAV subtypes. However, to date, a hydrolysis probe-based RT-PCR assay for the H5 marker in IAV has not been applied to wastewater.

In this study, we use prospective measurements of IAV M gene, which detects all IAV subtypes, in wastewater solids from 190 wastewater treatment plants (WWTPs) to identify WWTP showing springtime increases in IAV concentrations after the typical influenza season ends, coincident with the identification and spread of HPAI in animal populations in the US. We developed a novel RT-PCR hydrolysis-probe assay to detect the H5 subtype of IAV. After assay validation, we applied it to samples to measure concentrations of the H5 marker retrospectively from three wastewater treatment plants located near the apparent epicenter of the HPAI outbreak.

## Methods

### H5 assay design

Influenza A H5 subtype genome sequences were downloaded from the National Center for Biotechnology Information (NCBI) in Feb 2023 and supplemented with additional newer sequences available from NCBI and GISAID in April 2024. Sequences were aligned, and primers and probes were designed to target the hemagglutinin (HA) gene using Primer3Plus. Parameters used in assay development (eg, sequence length and GC content) are provided elsewhere^10^. The forward primer is: TATAGARGGAGGATGGCAGG, reverse primer: ACDGCCTCAAAYTGAGTGTT, and probe: AGGGGAGTGGKTACGCTGCRGAC (also in Table S1, Figure S1). The primers and probes were confirmed to be specific and sensitive for influenza A containing the H5 subtype of the HA gene *in silico* using NCBI BLAST.

The assay was tested *in vitro* against nucleic acids from a large collection of respiratory pathogens (Table S2) including non-H5 influenza subtypes. When panels were composed of whole viruses or bacteria, their nucleic-acids were extracted and purified as described below for the wastewater solids samples and then used neat as template in droplet digital RT-PCR assays. Assays were run in a single well using the cycling conditions and post processing using a droplet reader as described below for the wastewater samples in singleplex. Synthetic nucleic acid gene blocks purchased from IDT (Coralville, Iowa) were used as a positive control for the H5 marker.

### Prospective IAV M gene testing in wastewater

Wastewater samples were obtained from 190 wastewater treatment plants located in 41 states for a prospective wastewater monitoring project. Samples are collected 3 to 7 times per week between 7/1/23 and 4/22/24 (month/day/year format, n=19,323). Samples are shipped overnight to the laboratory at 4°C where they were processed immediately. During the time between sample collection and transport (0-3 days), we expect minimal losses of the RNA target based on results of IAV RNA persistence studies^18^. Upon receipt of the laboratory, samples are processed immediately to measure IAV M gene and PMMoV gene concentrations in the solid fraction of wastewater. Since the methods are described in detail in several different publications^8,19^, they are not provided in detail here. PMMoV is an endogenous plant virus present in wastewater and serves as an internal process control. Briefly, solids are isolated from the samples, suspended in a buffer at a low enough concentration so as to minimize inhibition^19^, and subjected to nucleic-acid extraction and purification (Chemagic Viral DNA/RNA 300 Kit H96, PerkinElmer, Shelton, CT), and inhibitor removal (Zymo OneStep PCR Inhibitor Removal Kit, Irvine, CA). Nucleic-acids from 6 to 10 aliquots of each sample are obtained and each used neat as template in 6 to 10 replicate droplet digital RT-PCR reaction wells to measure IAV and 1:100 diluted in 2 to 10 replicate wells to measure PMMoV. The process includes negative and positive extraction and PCR controls, as well as a measure of recovery as determined from spiking exogenous bovine coronavirus. The M gene target is present in all subtypes of IAV including the H5N1 subtype and the current circulating clade 2.3.4.4b. Concentrations are expressed as copies per gram dry weight. After obtaining these measurements, nucleic acids are biobanked at -80°C.

### Retrospective Influenza A M gene and H5 testing in wastewater

Biobanked samples between February 4 and April 16, 2024 from two wastewater treatment plant (WWTP) sites of the City of Amarillo Water Utilities Division in Texas (hereafter described as the “North” and “South” sites in Potter/Randall County, TX), as well as a WWTP in Dallas County, Texas were used to measure concentrations of H5 marker (samples were biobanked for up to 2 months, Table S3 provides exact dates of samples run, n=78 samples). The Potter/Randall County City of Amarillo plants provide 24-hour composited influent samples and the Dallas County plant provides settled solids from the primary clarifier. Plants service areas with separate sanitary sewer systems. The South and North sites in Potter/Randall County and the Dallas County site serve approximately 60,000, 140,000, and 186,000 people. Several industries are permitted to discharge wastewater to both City of Amarillo Water Utilities Division sanitary sewer systems including animal product processing facilities.

Concentrations of the H5 marker and the IAV M gene were measured in multiplex using an AutoDG Automated Droplet Generator (Bio-Rad, Hercules, CA), Mastercycler Pro (Eppendforf, Enfield, CT) thermocycler, and a QX600 Droplet Reader (Bio-Rad). The IAV M gene was measured again in the biobanked samples to document potential RNA degradation. In addition an assay for the N gene of SARS-CoV-2 was included (results not provided herein). Primers and probes for all the assays were purchased from IDT (Coralville, Iowa). The H5 probe contained the fluorescent molecule FAM and the IAV M gene probe contained ROX; both probes also contained ZEN, a proprietary internal quencher from IDT; and IBFQ, Iowa Black FQ. Each of the 6 replicate nucleic-acid extractions obtained from each sample were run neat as template in their own RT-PCR well to measure the RNA targets. The cycling conditions were as follows: reverse transcription at 50 °C for 60 min, enzyme activation at 95 °C for 5 min, 40 cycles of denaturation at 95 °C for 30 s and annealing and extension at 59 °C for 30 s, enzyme deactivation at 98 °C for 10 min then an indefinite hold at 4 °C. The ramp rate for temperature changes were set to 2 °C/second and the final hold at 4 °C was performed for a minimum of 30 min to allow the droplets to stabilize. Droplets were analyzed using the QX600 Droplet Reader (Bio-Rad). A well had to have over 10,000 droplets for inclusion in the analysis. All liquid transfers were performed using the Agilent Bravo (Agilent Technologies, Santa Clara, CA). Extraction and PCR positive and negative controls were run on each 96-well plate.

Results from replicate wells were merged for analysis. Concentrations of the targets are presented as copies per gram dry weight. Dry weight was determined using an aliquot of solids and drying in an oven^19^. For a sample to be scored as a positive, there had to be at least 3 positive droplets. The lowest measurable concentration is approximately 1000 copies/g dry weight. Errors are reported as standard deviations on the measurements as obtained from QX Manager Software (Bio-Rad, version 2.0).

### Influenza clinical data

Data on influenza-related emergency department visits by health district^20^ and statewide influenza positivity rates^21^ for Texas were obtained from publicly available data from the Texas Department of State Health Services. Potter/Randall County sites are within public health district 1, and the Dallas County site is within public health district 2/3.

### Data analysis

Across the 190 WWTPs in the prospective study, we identified onset and offset of IAV wastewater events using a previously published logic^16^. In short, IAV wastewater event onset occurred when all samples analyzed in the previous 14 days had concentrations of the IAV M gene ≥ 2000 cp/g and offset when less than or equal to 50% of the samples collected in the previous 14 days had concentrations ≥ 2000 cp/g. Using this approach, we identified WWTPs that showed a secondary onset of IAV wastewater events after the typical seasonal wastewater event offset. We also visually examined IAV/PMMoV concentrations at WWTPs to subjectively identify those with a noticeable spike, or increasing trends in concentrations observed after February 2024 when it was established that H5N1 was circulating.

## Results

### Assay sensitivity and specificity and QA/QC

Both *in silico* and *in vitro* testing indicated the H5 assay is 100% specific. *In vitro* tests returned non-detects for all non-target pathogens, including other influenza subtypes. The sensitivity of the assay is estimated to be 90% as 10% of the influenza A H5 subtype sequences in GISAID and NCBI have single nucleotide polymorphisms in the primer and probe regions. The H5 assay detects the circulating H5N1 clade 2.3.4.4b.

Results are reported following Environmental Microbiology Minimal Information^22^ (EMMI) guidelines (checklist available with all wastewater data at https://purl.stanford.edu/tr782wk3364). All positive and negative controls were positive and negative, respectively indicating no contamination. Nucleic-acid extraction efficiency for samples passed the quality control threshold of being over 0.1 for bovine coronavirus (median = 1.1, IQR = 0.8 - 1.5); recoveries greater than one are likely a result of errors associated with the measurements. PMMoV was detected at high concentrations in all samples (median of 3.3x10^8^, IQR = 1.9x10^8^-6.4x10^8^ cp/g) providing further support of efficient nucleic-acid extraction. We compared concentrations of IAV M gene in samples measured without any storage (during prospective monitoring) to those measured in the same biobanked samples; the median ratio of IAV M gene concentrations in biobanked to fresh samples was 1.3 (interquartile range 1.0 - 1.7, n=78) suggesting limited degradation of RNA during storage.

### Springtime increases in IAV M gene concentrations in WWTPs across the US

Using the prospectively measured IAV RNA concentrations, we found, using all data available on 24 April, 2024, that 44 of the 190 WWTPs showed a secondary IAV onset event after their seasonal IAV event offset (Table S4); these are located across 18 different states. There were 99 WWTPs for which the seasonal IAV wastewater event had not offset by the end of the data series; their IAV concentrations were visually examined for recent increases during March and April, and 15 locations (Table S3) were identified. Those 15 locations spanned 8 states. Example data from plants with distinct spring increases are shown in Figure S2.

### H5 marker prevalence in wastewater samples

Given that increases in wastewater appear to be coincident with the identification of avian influenza circulating in the nation (Figure S2), we decided to investigate whether the H5 marker was present in samples from three “sentinel” WWTPs located in Texas where H5N1 was identified in cows^14^ (Figure 1). We found that prior to early to mid-March (depending on the WWTP), the H5 marker was non-detect in the wastewater solids. Subsequently, as IAV M gene increased at the WWTPs, H5 marker became detectable at similar concentrations to the IAV M gene. When detected, H5 concentration ranged from 5847 to 2,158,821 (median = 29,364) cp/g. The IAV M gene concentrations measured during this time period are among the highest ever measured in wastewater (Figure S3). In samples where H5 was detected,the median ratio to IAV M gene was 0.6 at all three plants (IQR at Duck Creek: 0.6-0.6, Hollywood Road: 0.2 - 0.7, River Road: 0.4 - 0.8) During this time, influenza surveillance in Texas showed declined trends in influenza-related emergency department visits in the associated public health regions (Figure 2).

**Figure 1.**
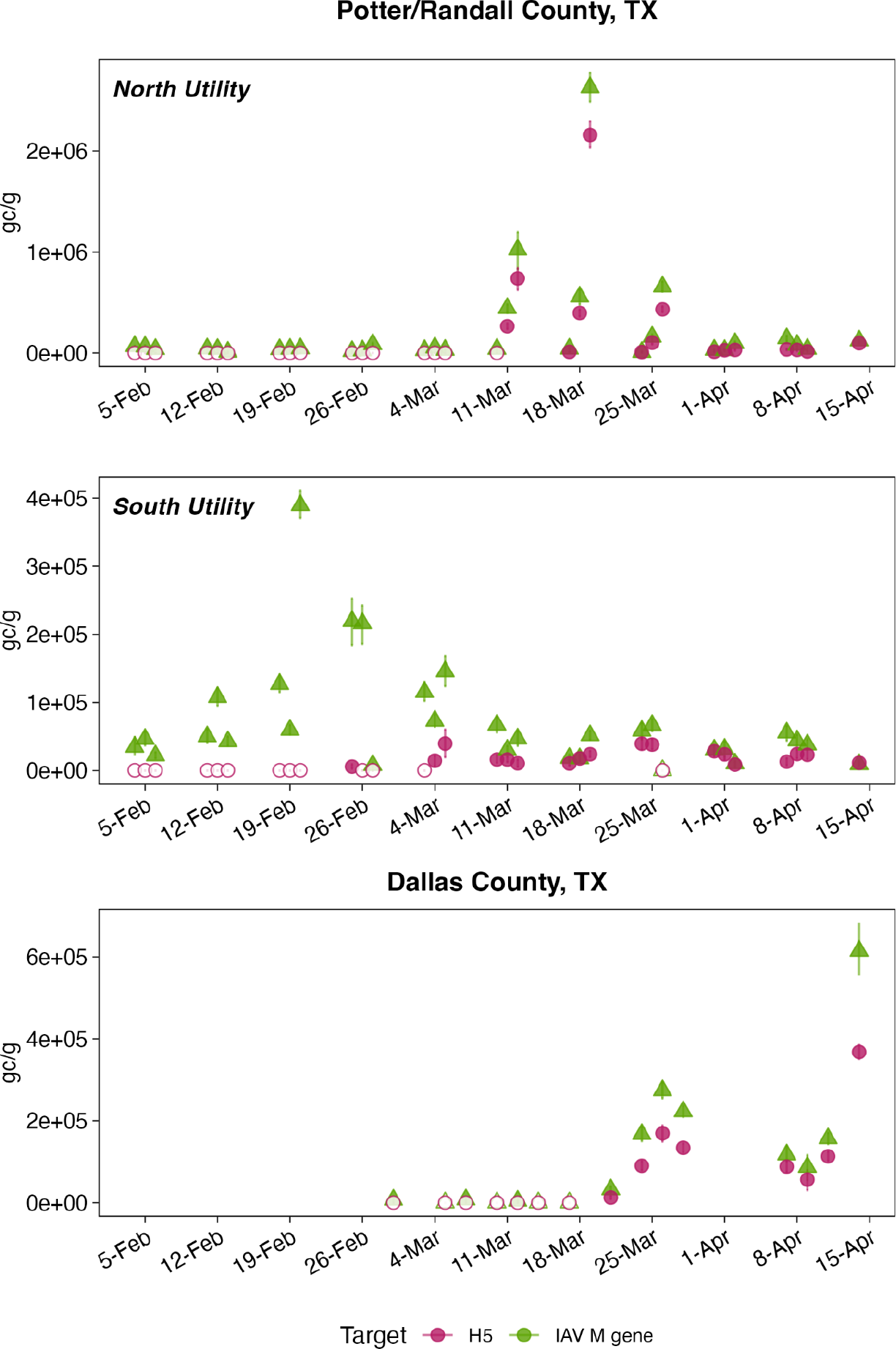
Concentrations of IAV M gene target (labeled as IAV, in green) as well as the H5 marker (labeled H5) and associated standard error in units of copies per gram dry weight; if error bars cannot be seen, then they are smaller than the symbols. Open markers are below the lowest measurable concentration (1000 copies per gram dry weight).

**Figure 2.**
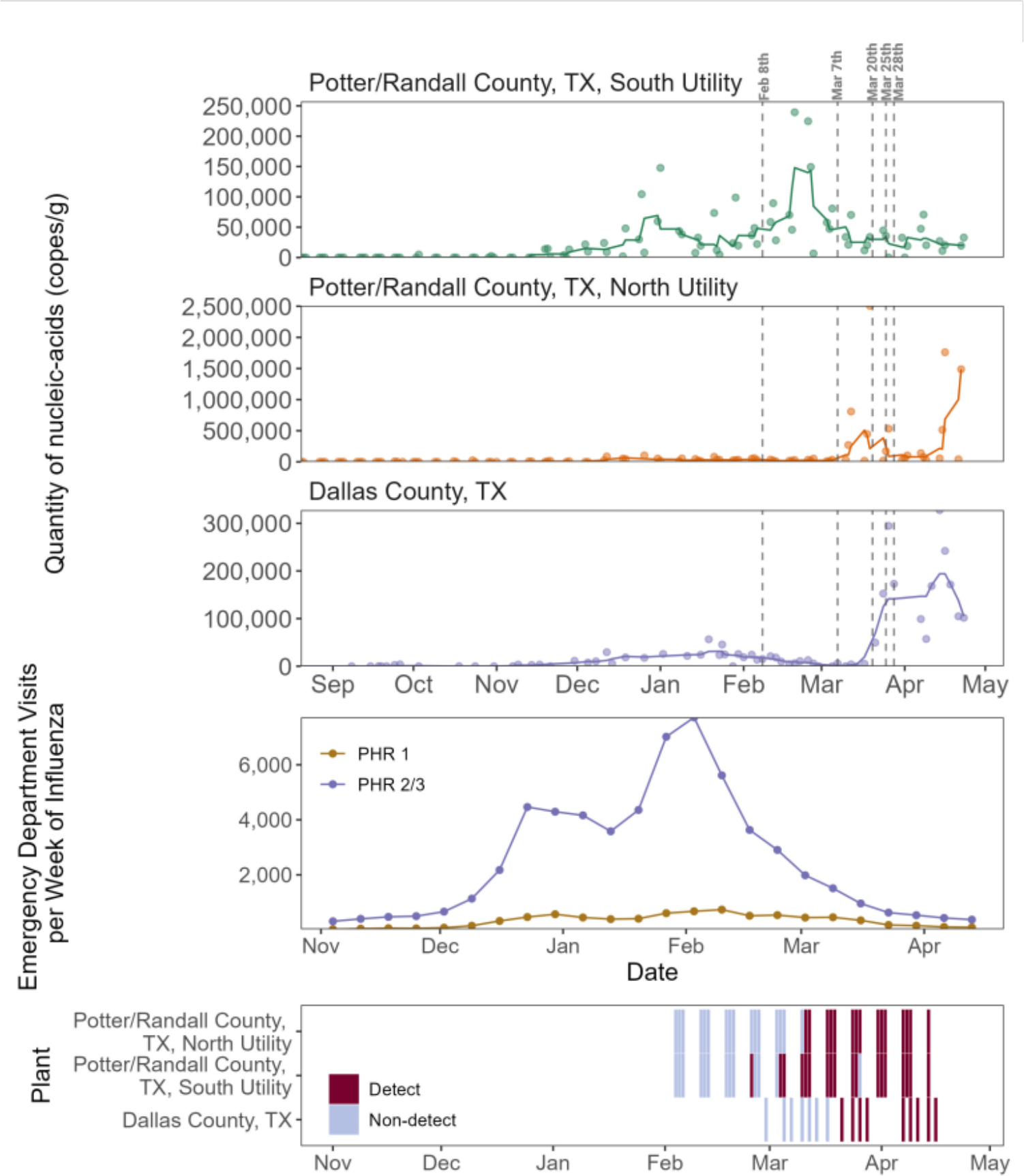
Top three panels. Concentrations of the IAV M gene in wastewater solids at the three WWTPs in Texas, as measured in the prospective monitoring. Line represents the 5-adjacent sample trimmed average, the symbols show the measured concentrations and the standard errors. Fourth panel from the top shows emergency department visits per week for influenza; PHR 1 represents Texas Public Health Region 1 (containing Potter/Randall Counties) and PHR2/3 represents Public Health Region 2/3 containing Dallas County. Bottom panel shows the presence or absence of the H5 marker in the samples from the three plants. Vertical lines indicate key dates as follows. February 8, 2024 USDA declares ongoing HPAI poultry outbreak, March 7, 2024: Unknown dairy cattle illness first reported, March 20, 2024: Samples collected from dairy cattle in Texas, March 25, 2024: Texas confirms H5N1 in dairy cattle, March 28, 2024: Positive H5N1 specimen collected from human.

## Discussion

H5 influenza RNA was readily detected in wastewater solids at sites with atypical increases in influenza A M gene concentrations in March and April 2024 during a period of a known H5N1 outbreak in the US. The presence of H5 in municipal wastewater indicates that contributions into the wastewater system include excretions or byproducts containing influenza viral RNA with an H5 subtype, but do not indicate the species that may be shedding an H5 influenza. During March and April 2024, increases in IAV M gene concentrations at these plants were not associated with increases in human influenza activity, as documented by the emergency room influenza case data in Texas. However, both the increases in M gene concentrations and H5 detection occurred immediately prior to and during reported outbreaks of H5N1 in dairy cattle in the region^14^. These results suggest that wastewater monitoring is a viable method of monitoring certain animal pathogens, and can provide a leading edge of detection that is of particular importance for diseases with zoonotic potential like HPAI.

### Hypothesized sources of H5

H5N1 outbreaks in dairy cattle were reported on March 25, 2024 in the Texas panhandle^23^, which contains the Potter/Randall County area. There was also a human case of H5N1 reported in Amarillo, TX, in an individual who worked at a commercial dairy cattle farm and reported close proximity to dairy cows^24^. As of April 25, 2024, no outbreaks have been reported within the vicinity of Sunnyvale, TX. Although no H5N1 outbreaks have been reported within the sewersheds of the WWTPs in this study, both City of Amarillo Water Utilities Division receive effluent from industries handling animal bioproducts, including dairies that receive milk from across the region and beef processing plants (personal communication from local staff). At least one dairy processing site is also permitted within the Dallas County sewershed area. Dairy cattle have been reported to shed H5N1, as well as other viruses, in milk in high quantities^25,26^. During the current outbreak, H5N1 has been detected in both raw and pasteurized milk, according to USDA^27^. Pasteurization is expected to inactivate the virus, although it may still be detectable as non-infectious genetic material can persist past pasteurization^28^. Regardless of whether milk or milk products entering the sewer system have undergone pasteurization or pre-treatment before they are discharged, our methods detect viral nucleic acids and their detection does not imply viability.

Given the outbreaks in dairy milking cattle in the region^14^, reports of H5N1 shedding in cow milk and detection in milk products on the shelf, and animal industry contributions into wastewater at the three utilities, we hypothesize that dairy processing discharge into the sewage system is driving the detection of H5 identified in wastewater solids. It is important to note that, even if true, this does not eliminate the possibility of contributions from other animals, including humans. If dairy industry activities in these sewersheds are a primary source of H5 in wastewater, this suggests that there are may be additional, unidentified outbreaks among cattle with milk sent to these facilities since milk from infected animals is required to be diverted from food supply.

### Limitations

The assay used here detects H5 broadly, and as such is not specific to H5N1 or HPAI, and may include other, low pathogenic H5 influenza viruses (although these are not expected to be circulating in these areas at this time^29^). HPAI genomes contain one copy each of M and HA genes, so results of the assays should be comparable, barring potential variability in assay performance and measurement errors. Caution should be taken in interpreting these results for characterizing the current H5N1 outbreak; without isolating possible sources in the sewer network of the H5 contributions our hypotheses about the source of the signal cannot be fully substantiated. However, multiple lines of evidence suggest animal sources.

This work shows that wastewater monitoring can provide an early warning for outbreaks likely to produce contributions to the sewershed outside of the expected human-associated inputs, including for animal outbreaks of diseases with zoonotic potential. In Potter/Randall County, retrospective testing shows that H5 was detectable in wastewater on Mar 1, 2024 - nearly a week before an unspecified disease was reported in dairy cattle in Texas (Mar 7) and several weeks before the causative agent was identified as H5N1 (Mar 25). Understanding and describing industry inputs into municipal wastewater, especially those that may be associated with contributions from animals, is critical both to identifying anomalies in influenza data from wastewater that do not relate to human disease, and identifying cases when wastewater can provide important, early warning data on outbreaks with zoonotic potential. Future work should further explore the possible contributions of animal shedding to municipal wastewater systems, and seek to sequence influenza A in wastewater to support genomic surveillance.

## Data Availability

All data produced are available online at the Stanford Digital Repository https://purl.stanford.edu/tr782wk3364.

## Acknowledgements

We acknowledge Todd Bell, Casie Stoughton, Jason Williams, Steve Kean, Richard Gibson, Tyler Edwards, Garry Singer, and Megan Sandefer for their assistance with the project.

## Data availability statement

All wastewater data are available at the Stanford Digital Repository: https://purl.stanford.edu/tr782wk3364.

## Supporting information

Additional methods, Figure S1, S2, and S3, and Tables S1, S2, S3, and S4.

