## Supporting information for "Detection of hemagglutinin H5 influenza A virus sequence in municipal wastewater solids at wastewater treatment plants with increases in influenza A in spring, 2024"

**Additional details related to EMMI guidelines.** The 78 wastewater samples from the study that were used for measuring H5 were selected at random for this analysis. The results for the samples from the prospective study are represented by results reported by Boehm et al.[1].

The average (standard deviation) number of partitions (droplets) for the across 6 replicate wells was 93902 (21281) for the multiplex reaction. The volume of the partitions, as reported by the machine vendor is 0.00085 μL. The average (standard deviation) of copies per partition for the H5 target is $5.0 \times 10^{-4}$ ($1.4 \times 10^{-3}$), and the average (standard deviation) of copies per partition for the IAV M gene target is $9.7 \times 10^{-4}$ ($1.9 \times 10^{-3}$).. An example fluorescent plot from the QX600 (6 color reader) is included in the Stanford Digital Repository with the deposited wastewater data (https://purl.stanford.edu/tr782wk3364).

**Figure S1. Location of the primers and probe within a reference genome (**NC_007362). Within the HA segment of A/goose/Guangdong/1/1996, ([https://www.ncbi.nlm.nih.gov/datasets/taxonomy/93838/](https://www.ncbi.nlm.nih.gov/datasets/taxonomy/93838/) ), the amplicon is located between bases 1086-1256. IAV_H5_P1 is the probe, IAV_H5_F1 and IAV_H5_R3 are the forward and reverse primers, respectively.

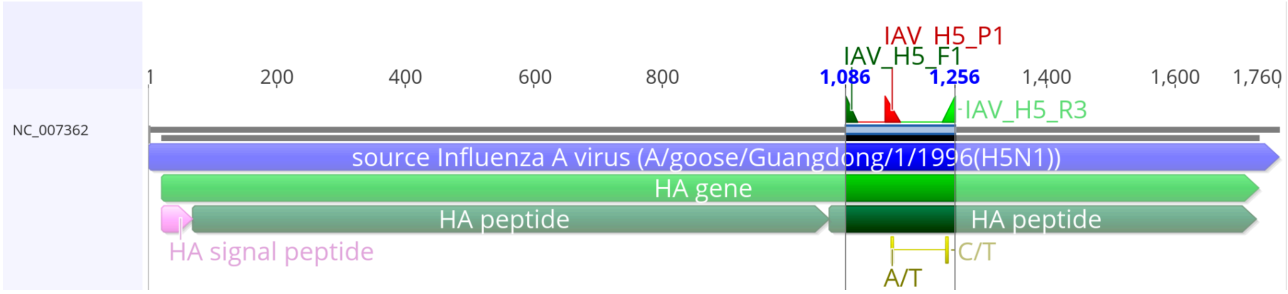

Figure S2. Concentrations of IAV M gene (units of copies per gram dry weight) measured at four plants since late August 2023. These plants were chosen because they show a clear sharp increase in IAV M gene concentrations after the typical influenza season period between November and February. Symbols represent measured concentrations of samples, and lines represent the 5-adjacent sample, centered, trimmed averages. Errors on the measurements are standard deviations. Any point at the top of a plot shown without error bars represents samples with concentrations larger than that axis limit. Vertical lines indicate key dates in the avian influenza outbreak. February 8, 2024 USDA declares ongoing HPAI poultry outbreak, March 7, 2024: Unknown dairy cattle illness first reported, March 20, 2024: Samples collected from dairy cattle in Texas, March 25, 2024: Texas confirms H5N1 in dairy cattle, March 28, 2024: Positive H5N1 specimen collected from human.

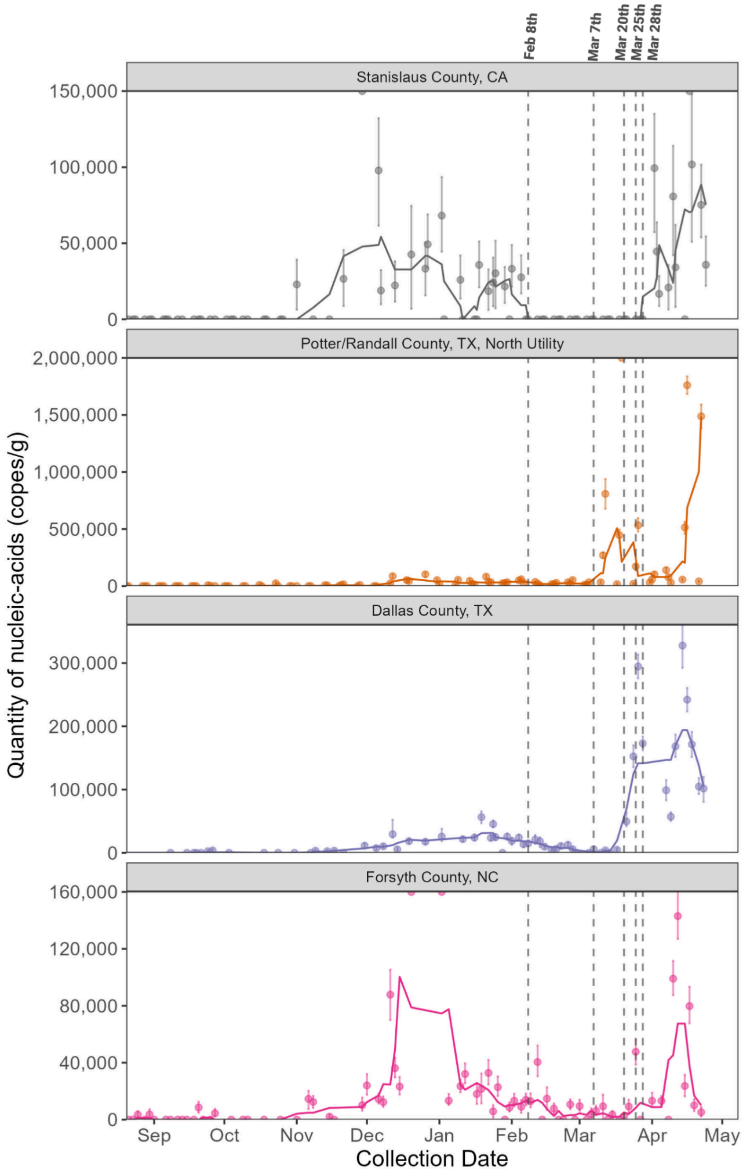

Figure S3. Histogram of IAV M gene concentrations from prospective monitoring described in the paper. The dashed lines indicate the maximum values of IAV M gene measurements made at the three WWTPs; these maximums occurred in March or April 2024.

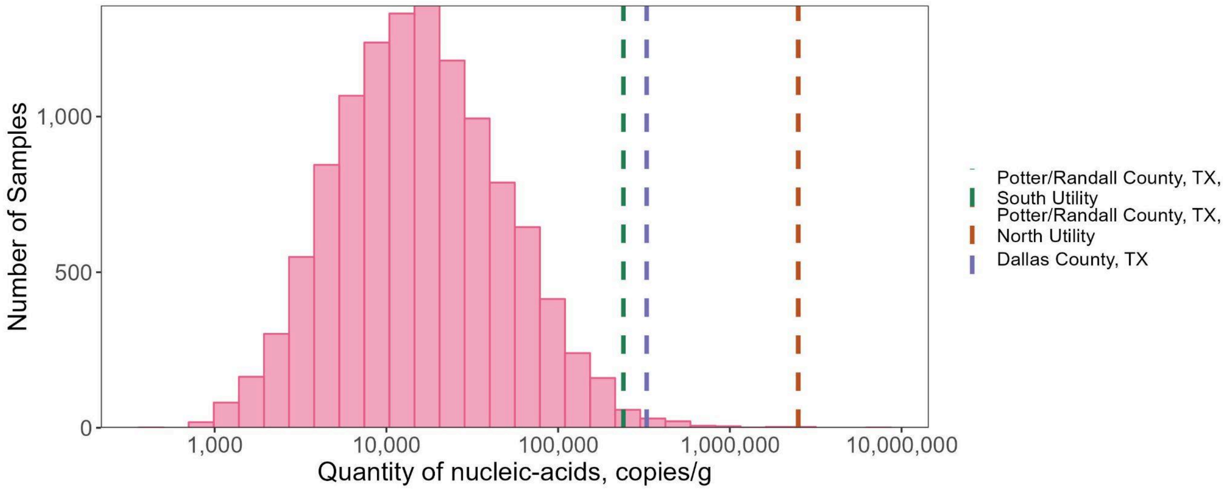

Table S1. Primers and probes developed to target the H5 version of the HA gene in influenza. The probe contained the fluorescent molecule fluorescein amidite (FAM) as well as ZEN, a proprietary internal quencher from IDT; and IBFQ, Iowa Black FQ.

| | |
|---|---|
| Forward primer | TATAGARGGAGGATGGCAGG |
| Reverse primer | ACDGCCTCAAAYTGAGTGTT |
| Probe | AGGGGAGTGGKTACGCTGCRGAC |

Table S2. Panel of pathogens used for in vitro specificity testing. All are inactivated pathogens from Zeptometrix (Buffalo, NY) unless “Twist” appears in front of the name in which case it is nucleic acid purchased from Twist (South San Francisco, CA).

| | | |
|---|---|---|
| Parainfluenza 1<br>Parainfluenza 2<br>Parainfluenza 3<br>Parainfluenza 4<br>Influenza A H1N1<br>Influenza AH1<br>Influenza AH3<br>Influenza B<br>Adenovirus 1<br>Adenovirus 3 | Adenovirus 31<br>Rhinovirus Type 1A<br>RSV A<br>RSV B<br>SARS-CoV-2<br>*M. pneumoniae*<br>*C. pneumoniae*<br>Metapneumovirus 8<br>Coronavirus HKU-1<br>Coronavirus 229E | Coronavirus NL63<br>Coronavirus OC43<br>*B. parapertussis*<br>*B. pertussis*<br>Twist Influenza B<br>Twist Influenza A H1N1<br>Twist Influenza A H3N2 |

Table S3. Sample date ranges for the three WWTPs. Dates are in month/day/year format.

| | Potter/Randall County, TX (North) | Potter/Randall County, TX (North) | Dallas County, TX |
|---|---|---|---|
| Date range | 2/4/24 - 4/14/24 | 2/4/24 - 4/14/24 | 2/29/24 - 4/16/24 |

Table S4. WWTPs with secondary IAV wastewater event onsets or increases (identified visually) in late March and early April, 2024.

| WWTPs with secondary springtime onset events | WWTPs still in seasonal onset with, visually identified recent increases |
|---|---|
| Bridgewater, NJ<br>Cahaba River, Birmingham, AL<br>Chattanooga, TN<br>East Bank, New Orleans, LA<br>Eastern, Orange County, FL<br>Esparto, CA<br>Fremont, CA<br>Gainesville, TX<br>Gautier, MS<br>Gilroy, CA<br>Kailua, Honolulu, HI<br>Kinston, NC<br>Lancaster, CA<br>Las Gallinas, San Rafael, CA<br>Little Falls Run, Stafford, VA<br>Lompoc, CA<br>Marina, CA<br>Modesto, CA<br>Muscatine, IA<br>Newark, CA<br>Northwest, Orange County, FL<br>Oakhurst, NJ<br>P20, Kansas City, KS<br>Pascagoula Moss Point, MS<br>Petaluma, CA<br>Regional, Laguna Niguel, CA<br>Riverside, CA<br>San Mateo, CA<br>Seaford, DE<br>Snohomish, WA<br>South River, Atlanta, GA<br>South, Laredo, TX<br>Sunnyvale, TX<br>University Park, PA<br>Village Creek, Birmingham, AL<br>Wahiawa, Honolulu, HI<br>West Bank, New Orleans, LA<br>West Contra Costa County, CA<br>West Railroad, San Rafael, CA<br>Wheeling, WV<br>Wichita Falls, TX<br>Windsor, CA<br>Woodland, CA<br>Zacate Creek, Laredo, TX | Bessemer, AL<br>Dallas Central, Dallas, TX<br>Hollywood Road, Amarillo, TX<br>Jenison, MI<br>Louisville, KY<br>Mill Valley, CA<br>Montpelier, VT<br>Sausalito, CA<br>River Road, Amarillo, TX<br>Salina, KS<br>South Bend, IN<br>Southside, Dallas, TX<br>St. Cloud, MN<br>White Rock Central, Dallas, TX<br>Wolcott, Kansas City, KS |
